# Features of 16,749 hospitalised UK patients with COVID-19 using the ISARIC WHO Clinical Characterisation Protocol

**DOI:** 10.1101/2020.04.23.20076042

**Authors:** AB Docherty, EM Harrison, CA Green, H Hardwick, R Pius, L Norman, KA Holden, JM Read, F Dondelinger, G Carson, L Merson, J Lee, D Plotkin, L Sigfrid, S Halpin, C Jackson, C Gamble, PW Horby, JS Nguyen-Van-Tam, ISARIC4C Investigators, J Dunning, PJM Openshaw, JK Baillie, MG Semple

**Author notes:** Corresponding Author and Guarantor, Prof. Malcolm G Semple PhD FRCPE FRCPHC FHEA, Institute in The Park, University of Liverpool, Alder Hey Children’s Hospital, Eaton Road, Liverpool, L12 2AP. ISARIC4C Investigators listed below. Annemarie B. Docherty, Centre for Medical Informatics, The Usher Institute, University of Edinburgh Ewen M Harrison, Centre for Medical Informatics, Usher Institute, University of Edinburgh Christopher A. Green, Institute of Microbiology & Infection, University of Birmingham Hayley Hardwick, NIHR Health Protection Research Unit in Emerging and Zoonotic Infections, and Institute of Infection and Global Health, Faculty of Health and Life Sciences, University of Liverpool. Riinu Pius, Centre for Medical Informatics, Usher Institute, University of Edinburgh Lisa Norman, Centre for Medical Informatics, Usher Institute, University of Edinburgh Karl A Holden, Institute of Translational Medicine, Faculty of Health and Life Sciences, University of Liverpool. Jonathan Read, Centre for Health Informatics, Computing and Statistics, Lancaster Medical School, Lancaster University, Bailrigg, United Kingdom LA1 4YG Frank Dondelinger, Centre for Health Informatics, Computing and Statistics, Lancaster Medical School, Lancaster University, Bailrigg, United Kingdom LA1 4YG Gail Carson, ISARIC Global Support Centre, Centre for Tropical Medicine and Global Health, Nuffield Department of Medicine, University of Oxford Laura Merson Infectious Diseases Data Observatory (IDDO), Centre for Tropical Medicine and Global Health, University of Oxford, UK Laura Merson ISARIC Global Support Centre, Centre for Tropical Medicine and Global Health, Nuffield Department of Medicine, University of Oxford James Lee, ISARIC Global Support Centre, Centre for Tropical Medicine and Global Health, Nuffield Department of Medicine, University of Oxford Daniel Plotkin, ISARIC Global Support Centre, Centre for Tropical Medicine and Global Health, Nuffield Department of Medicine, University of Oxford, UK Louise Sigfrid, ISARIC Global Support Centre, Centre for Tropical Medicine and Global Health, Nuffield Department of Medicine, University of Oxford Sophie Halpin, Liverpool Clinical Trials Centre, University of Liverpool Clare Jackson, Liverpool Clinical Trials Centre, University of Liverpool Carrol Gamble, Liverpool Clinical Trials Centre, University of Liverpool, Peter W. Horby, Centre for Tropical Medicine and International Health, Nuffield Department of Medicine, University of Oxford, Oxford, UK Jonathan S. Nguyen-Van-Tam, Division of Epidemiology and Public Health, University of Nottingham School of Medicine, Nottingham UK Jake Dunning National Infection Service, Public Health England, 61 Colindale Ave, London, NW9 5EQ and Faculty of Medicine, Imperial College London, Norfolk Place, London, W2 1PG Peter JM Openshaw National Heart and Lung Division, Faculty of Medicine, Imperial College London J Kenneth Baillie, Roslin Institute, University of Edinburgh, Edinburgh, UK and Intensive Care Unit, Royal Infirmary Edinburgh, UK Malcolm G Semple, NIHR Health Protection Research Unit in Emerging and Zoonotic Infections, and Institute of Translational Medicine, Faculty of Health and Life Sciences, University of Liverpool and Alder Hey Children’s Hospital, Liverpool. The Corresponding Author has the right to grant on behalf of all authors and does grant on behalf of all authors, an exclusive licence (or non-exclusive for government employees) on a worldwide basis to the BMJ Publishing Group Ltd to permit this article (if accepted) to be published in BMJ editions and any other BMJPGL products and sublicences such use and exploit all subsidiary rights, as set out in our licence. Prof. Semple affirms that the manuscript is an honest, accurate, and transparent account of the study being reported; that no important aspects of the study have been omitted; and that any discrepancies from the study as planned (and, if relevant, registered) have been explained.

## Abstract

**Objective:** To characterize the clinical features of patients with severe COVID-19 in the UK.

**Design:** Prospective observational cohort study with rapid data gathering and near real-time analysis, using a pre-approved questionnaire adopted by the WHO.

**Setting:** 166 UK hospitals between 6^th^ February and 18^th^ April 2020.

**Participants:** 16,749 people with COVID-19.

**Interventions:** No interventions were performed, but with consent samples were taken for research purposes. Many participants were co-enrolled in other interventional studies and clinical trials.

**Results:** The median age was 72 years [IQR 57, 82; range 0, 104], the median duration of symptoms before admission was 4 days [IQR 1,8] and the median duration of hospital stay was 7 days [IQR 4,12]. The commonest comorbidities were chronic cardiac disease (29%), uncomplicated diabetes (19%), non-asthmatic chronic pulmonary disease (19%) and asthma (14%); 47% had no documented reported comorbidity. Increased age and comorbidities including obesity were associated with a higher probability of mortality. Distinct clusters of symptoms were found: 1. respiratory (cough, sputum, sore throat, runny nose, ear pain, wheeze, and chest pain); 2. systemic (myalgia, joint pain and fatigue); 3. enteric (abdominal pain, vomiting and diarrhoea). Overall, 49% of patients were discharged alive, 33% have died and 17% continued to receive care at date of reporting. 17% required admission to High Dependency or Intensive Care Units; of these, 31% were discharged alive, 45% died and 24% continued to receive care at the reporting date. Of those receiving mechanical ventilation, 20% were discharged alive, 53% died and 27% remained in hospital.

**Conclusions:** We present the largest detailed description of COVID-19 in Europe, demonstrating the importance of pandemic preparedness and the need to maintain readiness to launch research studies in response to outbreaks.

**Trial documentation:** Available at https://isaric4c.net/protocols. Ethical approval in England and Wales (13/SC/0149), and Scotland (20/SS/0028). ISRCTN (pending).

## Introduction

In the wake of the A/H1N1pdm2009 influenza pandemic and the emergence of Middle East Respiratory Syndrome Coronavirus (MERS, 2012) it was recognised that the effectiveness of a response to a pandemic threat depended critically on the speed and focus of that response. The UK therefore set up and maintained a ‘sleeping’ pre-pandemic suite of documents, agreements and protocols in preparation for future outbreaks[1]. At the core of this plan was the Clinical Characterisation Protocol (CCP) for Severe Emerging Infection developed by the International Severe Acute Respiratory and emerging Infections Consortium (ISARIC).

ISARIC’s aim was to facilitate real-time research on diseases caused by novel pathogens of public health concern in order to save lives and inform public health policy early on and during outbreaks[2]. Our open-access protocols use standardised and refined case report forms, information and consent documents, and offer a tiered biological sampling schedule. This enables cross-correlation between sites and studies and allows comparisons to be made between diverse locations and treatment strategies[3].

ISARIC’s protocols were assigned Urgent Public Health Research Status by the National Institute for Health Research (NIHR), acknowledging that National Health Service (NHS) Hospitals in England and Wales and NIHR Clinical Research Network (CRN) resources would be necessary to prioritise their use in the event of activation. The ISARIC protocol gained approval across all 150 acute hospital trusts in England and Wales in 2013.

The World Health Organisation (WHO) Ethics Review Committee approved a global master protocol for the ISARIC CCP and endorsed its use in outbreaks of public health interest. The ISARIC CCP study is international and now includes low- and middle-income countries. The protocol is now known globally as the ISARIC-WHO Clinical Characterisation Protocol for Severe Emerging Infection and, in the UK, the ISARIC CCP-UK [ISRCTN pending CRN/CMPS ID:14152].

In response to the emergence and pandemic potential of SARS-CoV-2, the CCP-UK was activated on 17^th^ January 2020, in time to recruit the early patients of COVID-19 admitted to hospitals in England and Wales. Most people infected with SARS-CoV-2 have mild or moderate influenza-like illnesses with fever, cough and myalgia, but approximately one in five have illnesses of sufficient severity to warrant hospital admission.

ISARIC COVID-19 Clinical Characterisation Consortium (ISARIC4C) investigators have submitted regular reports to the UK Government’s New and Emerging Respiratory Virus Threats Advisory Group (NERVTAG). These reports are presented to the Scientific Pandemic Influenza Group on Modelling (SPI-M) and the Scientific Advisory Group for Emergencies ([SAGE](https://www.gov.uk/government/organisations/scientific-advisory-group-for-emergencies)). Aggregated data has been shared with WHO in the ISARIC COVID-19 report. We now present the first account of the clinical characteristics of patients from the ISARIC CCP-UK up to 18^th^ April 2020, and outcomes based on admissions on or before 6th April 2020.

## Methods

Clinical information from the routine health records of people admitted to 166 hospitals in England Wales and Scotland with proven SARS-CoV-2 infection were extracted into de-identified case report forms on a REDCap database (Research Electronic Data Capture, Vanderbilt University (USA) hosted by University of Oxford (UK), by 2,468 research nurses, administrators and volunteer medical students. With consent, additional biological samples were collected for research purposes.

Patient data were collected and uploaded from start of admission through to completion of the episode of care. Since the entry is not complete until the end of the episode, we chose *a priori* to restrict any analysis of outcome to patients who were admitted more than 14 days before data extraction (4^th^ April 2020). Data were analysed every 30 minutes using R (R Foundation for Statistical Computing, Vienna) and presented using RStudio Connect (RStudio, Boston) on a website to which UK public health agencies, government scientific advisory groups and health departments officials have secure access.

We have presented continuous variables as median [IQR] and categorical variables as n (%). We assessed the association of age with in-hospital mortality, adjusting for pre-existing patient characteristics (sex and comorbidity) using multivariable Cox proportional hazards regression.

The protocol, revision history, case report form, information leaflets and consent forms and detail of the Independent Data and Material Access Committee (IDAMAC) are available at https://isaric4c.net. The study was approved by the South Central - Oxford C Research Ethics Committee in England (Ref: 13/SC/0149), and by the Scotland A Research Ethics Committee (Ref: 20/SS/0028).

## Results

Between 6^th^ February and 14:00 on 18^th^ April 2020, CCP-UK recruited 16,749 patients admitted to hospital with COVID-19 in England, Scotland and Wales. This represents 14.7% of all people who have tested positive for COVID-19 in the UK, most of whom have not required hospital admission, and 28% of admissions with COVID-19.

The median time from onset of symptoms of COVID-19 in the community to presentation at hospital was 4 [IQR 1,8] days (n=9,785). The median duration of hospital stay was 7 [IQR 4,12] days.

A high proportion of patients required admission to High Dependency or Intensive Care Units 1,914/11,185 (17%).

### Age and Sex

This is a relatively elderly population, with median age 72 years [IQR 57, 82; range 0, 104] (figure 1). Only 239 patients (2.0%) are under 18 years and only 139 patients (1.1%) are under 5 years old. More men (60.2%, n=7,715) than women (39.8%, n=5,097) have been admitted to hospital with COVID-19 (missing data n=4,001). Fifty-five (6%) of women of reproductive age (n=963) are recorded as being pregnant.

**Figure 1:**
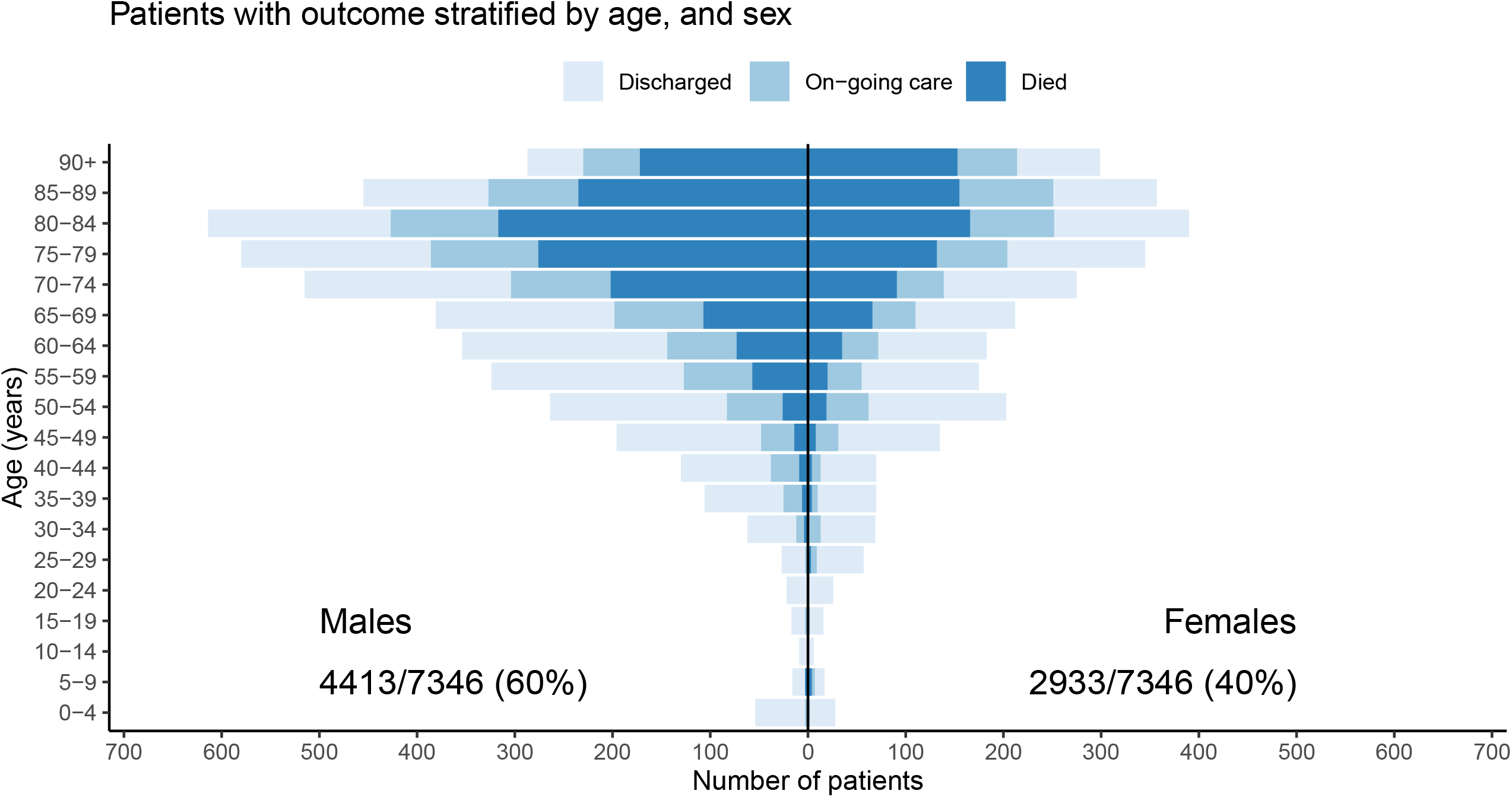
Patients with outcome (n=7,346) stratified by age, and sex. Bar fills are outcome (discharge/ongoing care/death) at the time of report (18/04/2020, n=16,749).

### Symptoms

The most common symptoms were cough (70%), fever (69%) and shortness of breath (65%) (figure 2A), though this reflects the case definition. Only 458 of 11,460 (4%) of patients reported no symptoms. There was a high degree of overlap between the three most common symptoms (figure 2B).

**Figure 2:**
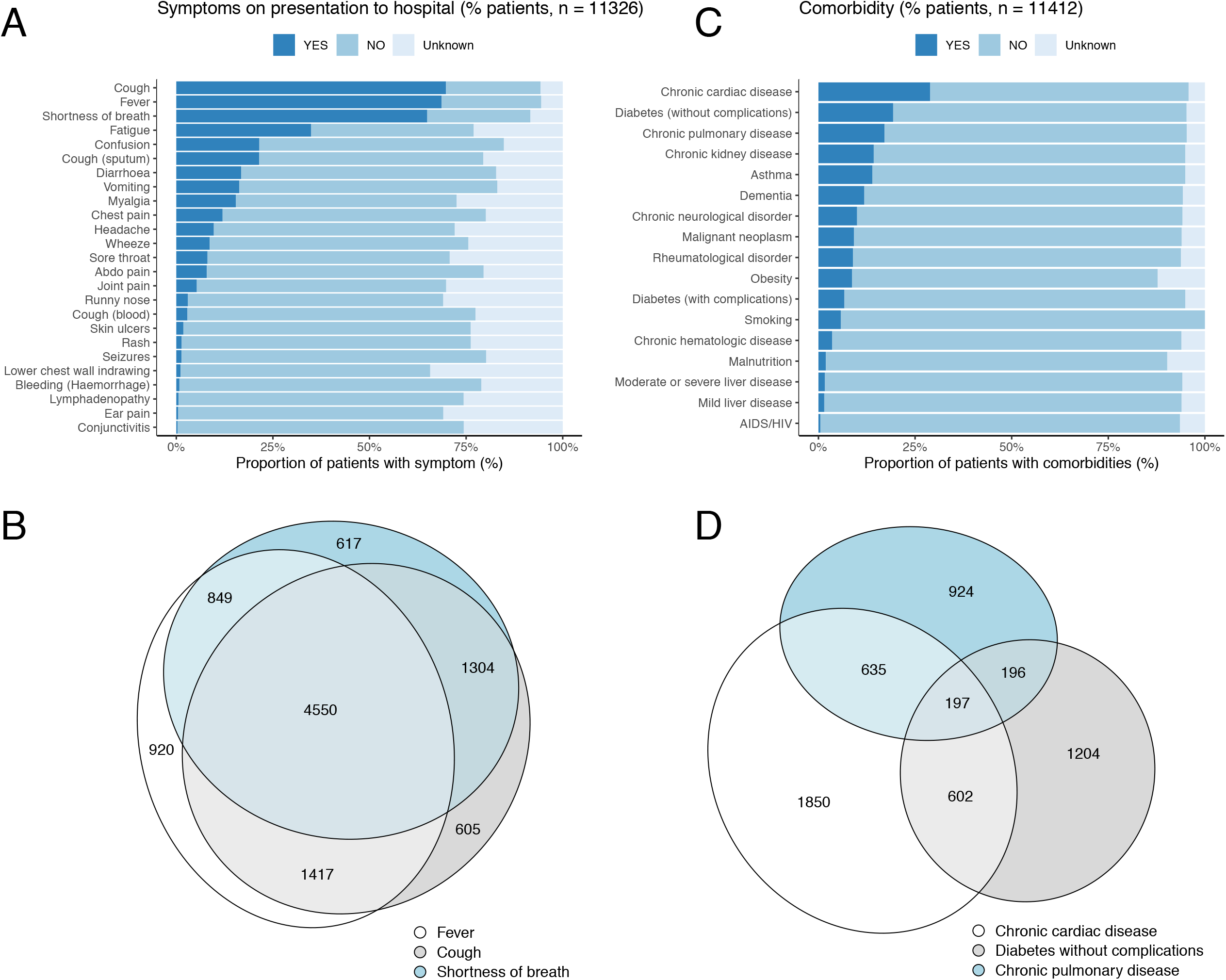
Presenting symptoms (n=11,326): A by frequency of presentation; B overlap of commonest symptoms. Comorbidities (n=11,412): C by frequency; D overlap of commonest comorbidities.

Clusters of symptoms on admission are apparent (figure 3). The most common symptom cluster encompass the respiratory system: cough, sputum, shortness of breath and fever. Two other clusters are also observed, one encompassing musculoskeletal symptoms: myalgia, joint pain and fatigue, and another of enteric symptoms: abdominal pain, vomiting and diarrhoea. 29% (3273/11460) of all patients complained of enteric symptoms on admission, mostly in association with respiratory symptoms, however 4% of all patients described enteric symptoms alone.

**Figure 3.**
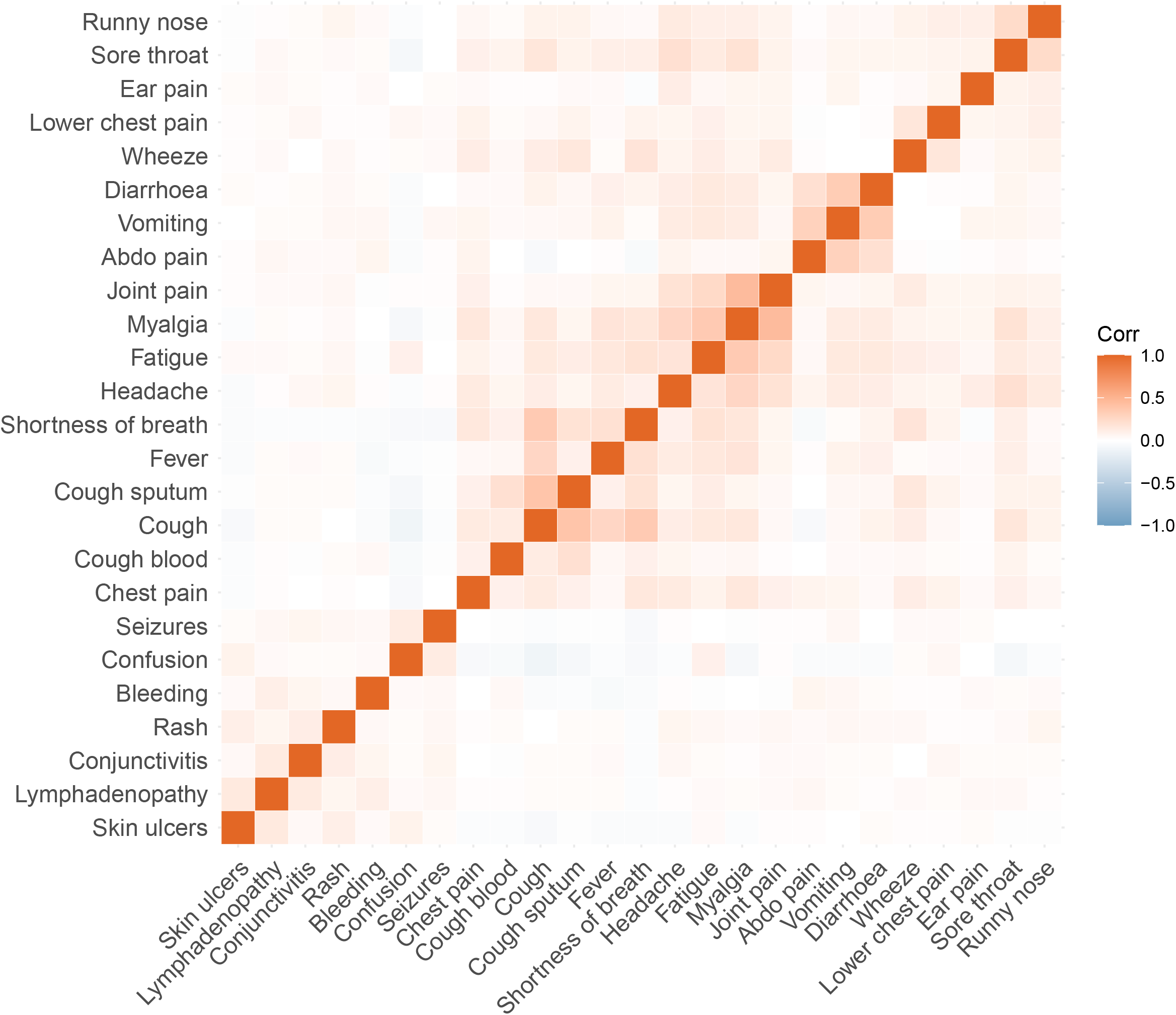
Correlation matrix of symptoms on admission to hospital.

### Comorbidities

Comorbidities at admission are shown in figure 2C. The most common recorded comorbidities are chronic cardiac disease (29%), uncomplicated diabetes (19%), chronic pulmonary disease excluding asthma (19%) and asthma (14%) (figure 2D). Of 16,749 patients, 7,924 (47%) patients had no documented reported comorbidity.

### Patient Outcomes

Status and outcomes for patients admitted at least 14 days before data extraction, were stratified by level of care (figure 4). Overall, 49% of patients were discharged alive, 33% have died and 17% continued to receive care at date of reporting. The median age of those who died in hospital from COVID-19 in the UK was 80 years, and only 12% of these patients had no documented comorbidity. For patients who received ward care, 55% were discharged alive, 31% have died and 14% remain in hospital. As expected, outcomes are worse for those who need higher levels of care. Of those admitted to critical care (intensive care/high dependency) 31% were discharged alive, 45% have died and 24% continued to receive care. Although the people receiving mechanical ventilation were younger than the overall cohort (61 y [IQR 52, 69]), only 20% had been discharged alive by 4^th^ April 2020, 53% have died and 27% are continuing to receive care. Duration of stay increased with age. Our decision to limit analysis of outcome for recent admissions to those admitted at least 14 days before date of reporting is supported by the description of proportion with known outcome by length of stay over time stratified by age (figure 5).

**Figure 4:**
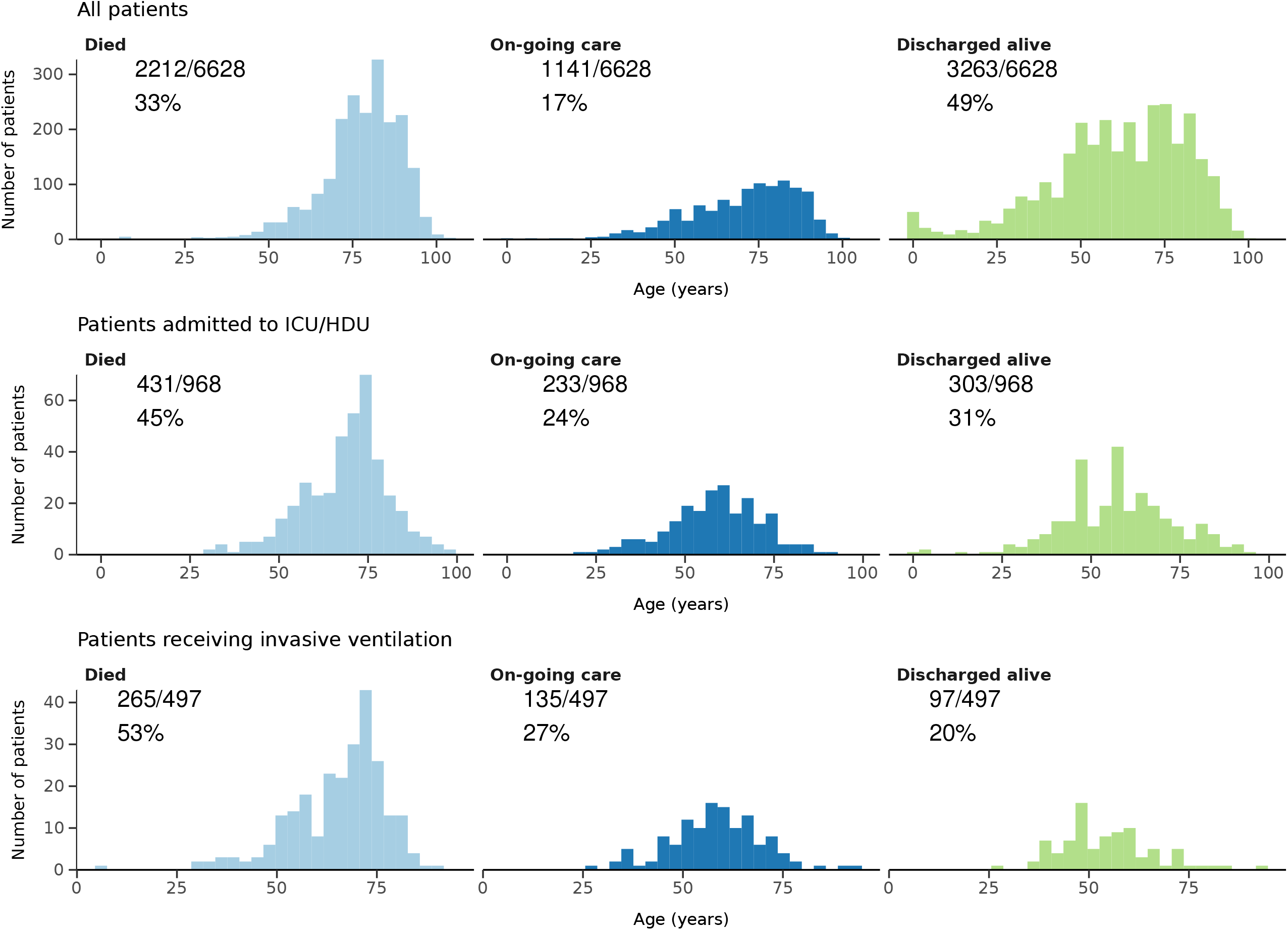
Status and outcome of patients admitted on or more than 14 days before reporting, stratified by level of care.

**Figure 5:**
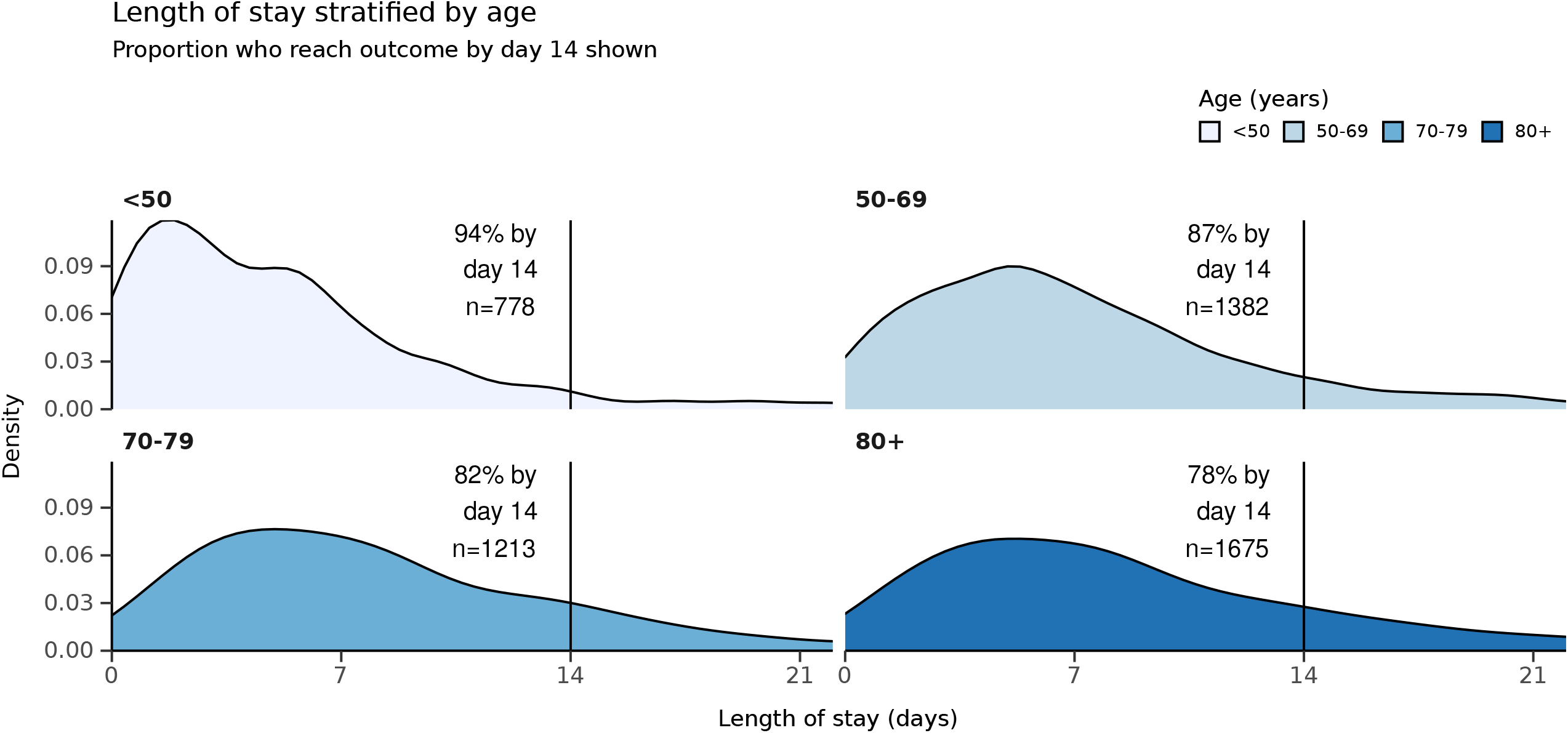
Length of stay stratified by age, with proportion reaching outcome at day 14 of admission: A all patients; B all ICU patients; C patients who have received invasive mechanical ventilation.

### Association of pre-existing patient characteristics and survival

Increased age was a strong predictor of in-hospital mortality after adjusting for comorbidity (figure 6): (reference age<50yrs) 50-69yrs Hazard ratio (HR) 4.02 (Confidence interval (CI) 2.88, 5.63, p<0.001), 70-79yrs HR 9.59 (CI 6.89, 13.34, p<0.001), >=80yrs HR 13.59 (CI 9.79, 18.85, p<0.001). Female sex was associated with lower mortality HR 0.80 (CI 0.72, 0.89, p<0.001). Comorbidities reported at admission were also associated with increased hospital mortality. This information must not be used as a predictive tool in practice or to inform individual treatment decisions.

**Figure 6:**
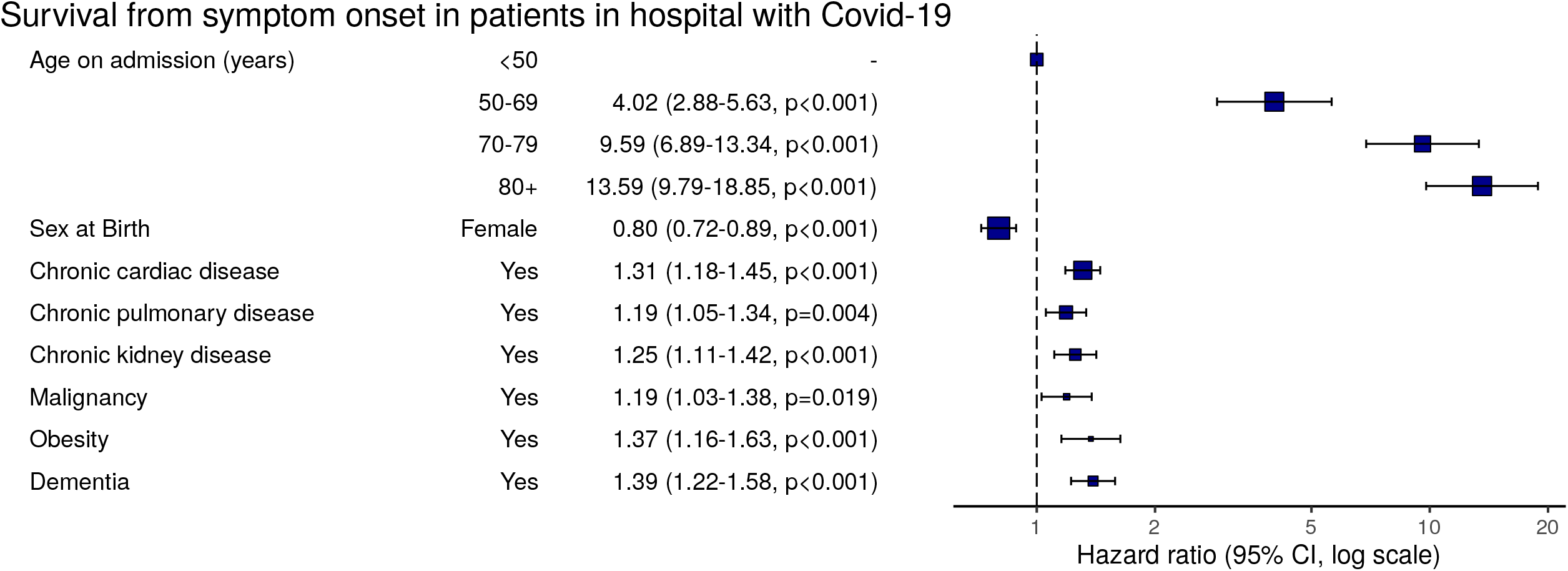
Multivariable Cox proportional hazards model (age, sex, comorbidities only) n=8341.

## Discussion

The ISARIC CCP-UK stood ready to activate large-scale studies of pandemic outbreaks for eight years enabling us to document patients in 166 hospitals across the UK in the early phase of the COVID-19 pandemic. This allowed a rapid and evidence-based response that had proved impossible in 2009 in the face of pandemic influenza. Studies such as this cannot be developed, approved and opened from the known start of a pandemic in time to inform case management and public health policy. Our study demonstrates the vital importance of forward planning and investment in preparedness.

The data presented here are the first description of patients during the growth phase of the SARS-CoV-2 pandemic in the UK. The first 101 patients reported were enrolled in the early phase of the outbreak as part of a High Consequence Infectious Disease containment strategy that ceased on 10^th^ March 2020. These and others were identified through screening further contact tracing in hospital and are represented in the 305 who were admitted without symptoms. The proportion of patients admitted to ICU in our study was higher than that in Italy at around 17%[4]. We are unable to capture treatment-limiting decisions regarding level of care.

The pattern of disease we describe broadly reflects that reported globally[5]. The current case definition of cough and fever, if strictly applied, would miss 7% of our hospitalised patients. A smaller proportion, 4% of patients, have presented with only enteric symptoms. This may be an underestimate since they fall outside standard criteria for testing. This enteric presentation risks misclassification of patients to non-COVID-19 care areas and may pose an additional nosocomial transmission risk. Severe SARS-CoV-2 infections are rare in those under 18 years of age, comprising only 1.4% of those admitted to hospital. Only 0.8% of those in our study were under 5 years of age. The “J” shaped age distribution is starkly different to the “U” shaped age distribution seen in seasonal influenza and “W” shaped distribution observed in the 2009 influenza pandemic [6]. It is not clear why SARS-CoV-19 has mostly spared children but we speculate this may be due to different expression of ACE2 receptor in the developing lung.

The recognition that obesity (as recognised by clinical staff) is associated with in-hospital mortality after adjustment for other comorbidities, age and sex has not been widely reported. Obesity was recognised as a risk factor in 2009 for pandemic A/H1N1 influenza, although not in 2016 Middle East respiratory syndrome coronavirus (MERS-CoV)[7,8].

There were few pregnant women in our cohort, the proportion (6%) being similar to the estimated proportion of pregnant women in the community[7]. Pregnancy was not associated with mortality, in apparent contrast with influenza[9].

The finding of independent associations of advancing age, male sex, chronic respiratory (though not asthma), chronic cardiac and chronic neurological disease with in-hospital mortality are in line with early international reports[10,11]. However, although age-adjusted mortality rates are high in the elderly, most of these patients were admitted to hospital with symptoms of COVID-19 and would not have died otherwise. It is notable that the enhanced severity in male patients is seen across all ages.

Mortality is high in patients admitted to general wards who are then not admitted to ICU, suggesting there is advanced care planning occurring between patients and physicians. Mortality rates are extremely high for those patients ventilated in ICU, compared with pandemic A/H1N1 influenza 2009 where ICU mortality was 31%[7]. Our data are in line with the initial ICNARC audit reports which represents ICUs in England, Wales and Northern Ireland[12]. Outcome analyses only included patients who were admitted before 4^th^ April to allow most patients to complete their journeys, however, there is an inherent reporting bias as the sickest of patients, particularly those admitted to ICU, have the longest hospital stays; in-hospital mortality rates may therefore increase. Also, as recruitment is occurring in the non-linear growth phase of the outbreak, recent patients for whom there are as yet no outcomes reported account for 18% of the total patients recruited.

While most patients with COVID-19 experience mild disease, of those who have been admitted to hospital 14 days prior to data extraction, half have been discharged alive and one third have died. Seventeen percent of those admitted to hospital required Critical Care. Those who have poor outcomes are more often elderly, male and obese.

The near-real time analysis of data presented by this Urgent Public Health study is allowing health policy makers to react dynamically to best evidence as it presents, such as expanded ICU capacity.

Our study is the first report in Europe of a very large and rapidly conducted study of COVID-19, demonstrating the vital importance of putting plans in place for the study of epidemic and pandemic threats and need to maintain them. It provides evidence of the pattern of disease in the UK population, identifies those sectors of the population at greatest risk and the use of healthcare resources. We welcome applications for data and material access via our Independent Data And Material Access Committee (https://isaric4c.net). Over the next few months we will issue reports in the BMJ on specific topics and analyses that are key to understanding the impact of COVID-19 and focusing on improving patient outcomes.

### Patient and Public Involvement

Patients or the public were not involved in the design, or conduct, or reporting, of this rapid response research.

## Data Availability

The protocol, revision history, case report form, information leaflets and consent forms and detail of the Independent Data and Material Access Committee (IDAMAC) are available at https://isaric4c.net.

https://isaric4c.net/protocols

## Acknowledgements

This work uses data provided by patients and collected by the NHS as part of their care and support #DataSavesLives. We are extremely grateful to the 2,648 frontline NHS clinical and research staff and volunteer medical students, who collected this data in challenging circumstances; and the generosity of the participants and their families for their individual contributions in these difficult times. We also acknowledge the support of Jeremy J Farrar, Nahoko Shindo, Devika Dixit, Nipunie Rajapakse, Piero Olliaro, Lyndsey Castle, Martha Buckley, Debbie Malden, Katherine Newell, Kwame O’Neill, Emmanuelle Denis, Claire Petersen, Scott Mullaney, Sue MacFarlane, Chris Jones, Nicole Maziere, Katie Bullock, Emily Cass, William Reynolds, Milton Ashworth, Ben Catterall, Louise Cooper, Terry Foster, Paul Matthew Ridley, Anthony Evans, Catherine Hartley, Chris Dunn, D. Sales, Diane Latawiec, Erwan Trochu, Eve Wilcock, Innocent Gerald Asiimwe, Isabel Garcia-Dorival, J. Eunice Zhang, Jack Pilgrim, Jane A Armstrong, Jordan J. Clark, Jordan Thomas, Katharine King, Katie Alexandra Ahmed, Krishanthi S Subramaniam, Lauren Lett, Laurence McEvoy, Libby van Tonder, Lucia Alicia Livoti, Nahida S Miah, Rebecca K. Shears, Rebecca Louise Jensen, Rebekah Penrice-Randal, Robyn Kiy, Samantha Leanne Barlow, Shadia Khandaker, Soeren Metelmann, Tessa Prince, Trevor R Jones, Benjamin Brennan, Agnieska Szemiel, Siddharth Bakshi, Daniella Lefteri, Maria Mancini, Julien Martinez, Angela Elliott, Joyce Mitchell, John McLauchlan, Aislynn Taggart, Oslem Dincarslan, Annette Lake.

## Potential Perceived Conflicts of Interest

AB Docherty reports grants from Department of Health and Social Care, during the conduct of the study; grants from Wellcome Trust, outside the submitted work; CA Green reports grants from DHSC National Institute of Health Research UK, during the conduct of the study; F Dondelinger is due to start a position at F. Hoffmann - La Roche on 4th May 2020; PW Horby reports grants from Wellcome Trust / Department for International Development / Bill and Melinda Gates Foundation, grants from NIHR, during the conduct of the study; JS Nguyen-Van-Tam reports grants from Department of Health and Social Care, England, during the conduct of the study; and is seconded to the Department of Health and Social Care, England (DHSC); PJM Openshaw reports personal fees from Consultancy, grants from MRC, grants from EU Grant, grants from NIHR Biomedical Research Centre, grants from MRC/GSK, grants from Wellcome Trust, grants from NIHR (HPRU), grants from NIHR Senior Investigator, personal fees from European Respiratory Society, grants from MRC Global Challenge Research Fund, outside the submitted work; and The role of President of the British Society for Immunology was an unpaid appointment but travel and accommodation at some meetings is provided by the Society; JK Baille reports grants from Medical Research Council UK; MG Semple reports grants from DHSC National Institute of Health Research UK, grants from Medical Research Council UK, grants from Health Protection Research Unit in Emerging & Zoonotic Infections, University of Liverpool, during the conduct of the study; other from Integrum Scientific LLC, Greensboro, NC, USA, outside the submitted work.

EM Harrison, H Ardwick, J Dunning, R Pius, L Norman, KA Holden, JM Read, G Carson, L Merson, J Lee, D Plotkin, L Sigfred, S Halpin, C Jackson, C Gamble, have nothing to declare.

## ISARIC Coronavirus Clinical Characterisation Consortium (ISARIC4C) Investigators

Consortium Lead Investigator J Kenneth Baillie, Chief Investigator Malcolm G Semple. Co-Lead Investigator Peter JM Openshaw. ISARIC Clinical Coordinator Gail Carson. Co-Investigators: Beatrice Alex, Benjamin Bach, Wendy S Barclay, Debby Bogaert, Meera Chand, Graham S Cooke, Annemarie B Docherty, Jake Dunning, Ana da Silva Filipe, Tom Fletcher, Christopher A Green, Julian A Hiscox, Antonia Ying Wai Ho, Peter W Horby, Samreen Ijaz, Saye Khoo, Paul Klenerman, Andrew Law, Wei Shen Lim, Alexander, J Mentzer, Laura Merson, Alison M Meynert, Mahdad Noursadeghi, Shona C Moore, Massimo Palmarini, William A Paxton, Georgios Pollakis, Nicholas Price, Andrew Rambaut, David L Robertson, Clark D Russell, Vanessa Sancho-Shimizu, Tom Solomon, Shiranee Sriskandan, David Stuart, Charlotte Summers, Richard S Tedder, Emma C Thomson, Ryan S Thwaites, Lance CW Turtle, Maria Zambon. Project Managers Hayley Hardwick, Chloe Donohue, Jane Ewins, Wilna Oosthuyzen. Data Analysts: Lisa Norman, Riinu Pius, Tom M Drake, Cameron J Fairfield, Stephen Knight, Kenneth A Mclean, Derek Murphy, Catherine A Shaw. Data and Information System Manager: Jo Dalton, Michelle Girvan, Egle Saviciute, Stephanie Roberts Janet Harrison, Laura Marsh, Marie Connor. Data integration and presentation: Gary Leeming, Andrew Law, Ross Hendry. Material Management: William Greenhalf, Victoria Shaw, Sarah McDonald. Local Principal Investigators: Kayode Adeniji, Daniel Agranoff, Ken Agwuh, Dhiraj Ail, Ana Alegria, Brian Angus, Abdul Ashish, Dougal Atkinson, Shahedal Bari, Gavin Barlow, Stella Barnass, Nicholas Barrett, Christopher Bassford, David Baxter, Michael Beadsworth, John Berridge, Nicola Best, Pieter Bothma, David Brealey, Robin Brittain-Long, Naomi Bulteel, Tom Burden, Andrew Burtenshaw, Vikki Caruth, David Chadwick, Duncan Chambler, Nigel Chee, Jenny Child, Srikanth Chukkambotla, Tom Clark, Paul Collini, Graham Cooke, Catherine Cosgrove, Jason Cupitt, Maria-Teresa Cutino-Moguel, Paul Dark, Chris Dawson, Samir Dervisevic, Phil Donnison, Sam Douthwaite, Ingrid DuRand, Ahilanadan Dushianthan, Tristan Dyer, Cariad Evans, Chi Eziefula, Chrisopher Fegan, Duncan Fullerton, Sanjeev Garg, Sanjeev Garg, Atul Garg, Jo Godden, Arthur Goldsmith, Clive Graham, Elaine Hardy, Stuart Hartshorn, Daniel Harvey, Peter Havalda, Daniel B Hawcutt, Antonia Ho, Maria Hobrok, Luke Hodgson, Anita Holme, Anil Hormis, Michael Jacobs, Susan Jain, Paul Jennings, Agilan Kaliappan, Vidya Kasipandian, Stephen Kegg, Michael Kelsey, Jason Kendall, Caroline Kerrison, Ian Kerslake, Oliver Koch, Gouri Koduri, George Koshy, Shondipon Laha, Susan Larkin, Tamas Leiner, Patrick Lillie, James Limb, Vanessa Linnett, Jeff Little, Michael MacMahon, Emily MacNaughton, Ravish Mankregod, Huw Masson, Elijah Matovu, Katherine McCullough, Ruth McEwen, Manjula Meda, Gary Mills, Jane Minton, Mariyam Mirfenderesky, Kavya Mohandas, James Moon, Elinoor Moore, Patrick Morgan, Craig Morris, Katherine Mortimore, Samuel Moses, Mbiye Mpenge, Rohinton Mulla, Michael Murphy, Thapas Nagarajan, Mark Nelson, Igor Otahal, Mark Pais, Selva Panchatsharam, Hassan Paraiso, Brij Patel, Justin Pepperell, Mark Peters, Mandeep Phull, Stefania Pintus, Jagtur Singh Pooni, Frank Post, David Price, Rachel Prout, Nikolas Rae, Henrik Reschreiter, Tim Reynolds, Neil Richardson, Mark Roberts, Devender Roberts, Alistair Rose, Guy Rousseau, Brendan Ryan, Taranprit Saluja, Aarti Shah, Prad Shanmuga, Anil Sharma, Anna Shawcross, Jeremy Sizer, Richard Smith, Catherine Snelson, Nick Spittle, Nikki Staines, Tom Stambach, Richard Stewart, Pradeep Subudhi, Tamas Szakmany, Kate Tatham, Jo Thomas, Chris Thompson, Robert Thompson, Ascanio Tridente, Darell Tupper - Carey, Mary Twagira, Andrew Ustianowski, Nick Vallotton, Lisa Vincent-Smith, Shico Visuvanathan, Alan Vuylsteke, Sam Waddy, Rachel Wake, Andrew Walden, Tony Whitehouse, Paul Whittaker, Ashley Whittington, Meme Wijesinghe, Martin Williams, Lawrence Wilson, Sarah Wilson, Stephen Winchester, Martin Wiselka, Adam Wolverson, Daniel G Wooton, Andrew Workman, Bryan Yates, Peter Young.

## References

1 Simpson CR, Beever D, Challen K, et al. The UK’s pandemic influenza research portfolio: a model for future research on emerging infections. Lancet Infect Dis 2019;19:e295–300.http://www.ncbi.nlm.nih.gov/pubmed/31006605 x(accessed 25 Sep 2019).

2 Dunning JW, Merson L, Rohde GGU, et al. Open source clinical science for emerging infections. Lancet Infect. Dis. 2014;14:8–9. doi:10.1016/S1473-3099(13)70327-X

3 Pardinaz-Solis R, Longuere K-S, Moore S, et al. ISARIC – enhancing the clinical research response to epidemics. Int J Infect Dis 2016;53:137. doi:10.1016/j.ijid.2016.11.338

4 Remuzzi A, Remuzzi G. COVID-19 and Italy: what next? Lancet. 2020;395:1225–8. doi:10.1016/S0140-6736(20)30627-9

5 Huang C, Wang Y, Li X, et al. Clinical features of patients infected with 2019 novel coronavirus in Wuhan, China. Lancet 2020;395:497–506. doi:10.1016/S0140-6736(20)30183-5

6 Myles PR, Semple MG, Lim WS, et al. Predictors of clinical outcome in a national hospitalised cohort across both waves of the influenza A/H1N1 pandemic 2009-2010 in the UK. Thorax 2012;67:709–17. doi:10.1136/thoraxjnl-2011-200266

7 Nguyen-Van-Tam Js, Openshaw PJM, Hashim A, et al. Risk factors for hospitalisation and poor outcome with pandemic A/H1N1 influenza: United Kingdom first wave (May-September 2009). Thorax 2010;65:645–51. doi:10.1136/thx.2010.135210

8 Alraddadi BM, Watson JT, Almarashi A, et al. Risk factors for primary middle east respiratory syndrome coronavirus illness in humans, Saudi Arabia, 2014. Emerg Infect Dis Published Online First: 2016. doi:10.3201/eid2201.151340

9 Clohisey S, Baillie JK. Host susceptibility to severe influenza A virus infection. Crit. Care. 2019;23. doi:10.1186/s13054-019-2566-7

10 Chen N, Zhou M, Dong X, et al. Epidemiological and clinical characteristics of 99 cases of 2019 novel coronavirus pneumonia in Wuhan, China: a descriptive study. Lancet Published Online First: 2020. doi:10.1016/S0140-6736(20)30211-7

11 Onder G, Rezza G, Brusaferro S. Case-Fatality Rate and Characteristics of Patients Dying in Relation to COVID-19 in Italy. JAMA - J. Am. Med. Assoc. 2020. doi:10.1001/jama.2020.4683

12 ICNARC. ICNARC report on COVID-19 in critical care. 2020;:1–9.https://www.icnarc.org/Our-Audit/Audits/Cmp/Reports (accessed 13 Apr 2020).

